# Causal modeling in large-scale data to improve identification of adults at risk for combined and common variable immunodeficiencies

**DOI:** 10.1101/2024.08.08.24311672

**Authors:** Giorgos Papanastasiou, Marco Scutari, Raffi Tachdjian, Vivian Hernandez-Trujillo, Jason Raasch, Kaylyn Billmeyer, Nikolay V Vasilyev, Vladimir Ivanov

## Abstract

Combined immunodeficiencies (CID) and common variable immunodeficiencies (CVID), prevalent yet substantially underdiagnosed primary immunodeficiency disorders, necessitate improved early detection strategies. Leveraging large-scale electronic health record (EHR) data from four nationwide US cohorts, we developed a novel causal Bayesian Network (BN) model to unravel the complex interplay of antecedent clinical phenotypes associated with CID/CVID. Consensus directed acyclic graphs (DAGs) were constructed, which demonstrated robust predictive performance (ROC AUC in unseen data within each cohort ranged from 0.77-0.61) and generalizability (ROC AUC across all unseen cohort evaluations ranged from 0.72-0.56) in identifying CID/CVID across diverse patient populations, created using different inclusion criteria. These consensus DAGs elucidate causal relationships between comorbidities preceding CID/CVID diagnosis, including autoimmune and blood disorders, lymphomas, organ damage or inflammation, respiratory conditions, genetic anomalies, recurrent infections, and allergies. Further evaluation through causal inference and by expert clinical immunologists substantiates the clinical relevance of the identified phenotypic trajectories within the consensus DAGs. These findings hold promise for translation into improved clinical practice, potentially leading to earlier identification and intervention for adults at risk of CID/CVID.

## Introduction

Primary immunodeficiencies (PI) are heterogeneous genetic disorders characterized by immune system defects [1]. PI patients are susceptible to life-threatening infections, malignancies, organ damage, severe allergies, and autoimmunity [2,3]. As of 2022, research has linked 485 PI phenotypes to 511 genetic defects [4,5] and this number is expected to increase with ongoing PI research [4–6].

PI is more common than originally thought. Recent studies suggest that PI affects 1-2% of the global population, with 70-90% of patients remaining undiagnosed [7, 8]. Early PI diagnosis is important to improve health outcomes but is hampered by the heterogeneous clinical presentation and low awareness among primary care practitioners leading to a lack of timely referrals [1–7, 9]. Misdiagnosis, underdiagnosis or diagnosis delay are therefore common in PI [1,2,7–10]. Undiagnosis is associated with increased mortality, morbidity, healthcare visits and costs [8–10]. Therefore, robust methods for systematic PI screening are urgently needed [1–5].

Combined immunodeficiencies (CID) are a subgroup of PI defined by both cellular (T-cell) and humoral (B-cell) immunity defects [1,8]. Common variable immunodeficiencies (CVID) are characterized by humoral immunity and are among the most frequent PI [1,2]. Severe CID (SCID), characterized by profound T-cell impairment, is life-threatening without early infancy treatment via newborn screening and bone marrow (BMT) or hematopoietic stem cell transplantation (HSCT) [1,11]. CID, excluding SCID, are marked by partial T-cell dysfunction, are associated with variable disease progression and are among the least investigated PI [1–8]. Unlike SCID, CID patients typically present with late symptom onset (>1-year of age) due to residual T-cell function [8]. Beyond SCID, there is no population-based screening method for PI, leading to many CVID/CID diagnoses only in adulthood due to delayed disease onset and lack of awareness hindering childhood diagnosis [1–9]. Despite the availability of definitive treatments like HSCT, BMT, and Ig replacement therapy [1, 8, 9], the lack of population-wide screening beyond SCID necessitates a systematic approach to identify at-risk adults, facilitating early referral and intervention [8, 9, 12].

Our work aimed to unravel the interplay between clinical diagnosis codes linked to CID/CVID through the development of a Bayesian Network (BN) model [16, 17]. Our recently developed machine learning (ML) model accurately identified CID/CVID from large-scale, nationwide (US) electronic health record (EHR) diagnosis codes, the same patient populations utilized in the present study [13]. Through descriptive statistical analysis, we further elucidated combinations of antecedent phenotypes correlated with CID/CVID [13]. Another study used ML on diagnosis codes from small-scale EHR to identify PI [14]. However, it is known that typical (non-causal) ML and statistical models are unaware of how the existence of causal relationships between variables can affect the overall reliability (generalizability, robustness, interpretability) of their outcomes [15]. Prior ML research has not prioritized identifying causal relationships and confounding variables, potentially limiting the generalizability, robustness, interpretability and clinical applicability of PI study outcomes. Addressing these factors could improve the early detection of PI, through the identification of causal paths in patient clinical history. A BN is a probabilistic graphical model that represents variables and their conditional dependencies via a directed acyclic graph (DAG) [16, 17]. A DAG can be learned from the data: its nodes represent data variables (e.g., diagnosis codes) with arcs indicating probabilistic dependencies [16]. Judea Pearl imbued BNs with causal semantics by interpreting them as causal networks [16]. By positing certain assumptions such as the absence of unobserved confounders, he established that arcs within a BN can be construed as representing direct causal relationships, enabling the identification and estimation of causal effects. In the context of PI diagnosis codes, a DAG can be used to identify clinical history traits: causal trajectories of clinical phenotypes associated with CID/CVID diagnosis. Since BN is a generative model, a DAG can subsequently be used to predict CID/CVID [16, 17]. Causal modeling can potentially improve the generalizability, robustness and interpretability of ML models [16–20]. While randomized clinical trials are the reference standard for establishing causal effects, they commonly face ethical, scalability, and patient disruption challenges [20]. EHRs serve as a rich source of real-world observational data, often providing the only accessible information for research purposes [13, 14]. Since we cannot directly randomize interventions with observational data, causal modeling relies on careful assumptions to account for potential biases and confounding factors [16, 17, 20]. Although BN-derived DAGs have been applied to real-world observational data in other biomedical fields, e.g., to identify genetic and protein interactions [18, 19], there is no previous work in the context of patient clinical history, i.e., identifying phenotypic trajectories and assessing their causal impact on CID/CVID diagnosis.

Leveraging large-scale observational EHR data from four nationwide US cohorts, we developed and evaluated causal BN models to elucidate the complex interplay of antecedent clinical phenotypes associated with CID/CVID. An ensemble approach was employed, constructing multiple BN models on bootstrapped datasets. Each resulting DAG was subsequently integrated into a consensus DAG, wherein arcs exhibiting lower prevalence across the ensemble were pruned. The consensus DAGs demonstrated robust predictive performance and generalizability in identifying CID/CVID patients, across diverse populations. These DAGs elucidate causal trajectories of interrelated comorbidities preceding CID/CVID diagnosis, including autoimmune and blood disorders, lymphomas, organ damage or inflammation, respiratory conditions, genetic anomalies, recurrent infections, and allergies. Causal inference analysis, quantifying the impact of each variable in the consensus DAG on the odds of receiving a CID/CVID diagnosis, and evaluations by expert clinical immunologists, substantiate further the clinical relevance of the identified phenotypic trajectories which hold promise for translation into refined clinical practices.

## Results

The study comprised four parts as follows: (1) A consensus DAG was learned for each cohort (Cohorts 1-4) and its predictive ability was evaluated using cross-validation. (2) The generalizability of each consensus DAG was then evaluated, by assessing their predictive accuracy in the other three unseen cohorts. (3) Quantitative assessments were conducted by performing causal interventions to evaluate the impact of each DAG variable on the CID/CVID diagnosis. (4) The transferability of these DAGs to clinical practice was assessed through qualitative evaluations with domain experts. Figure 1 illustrates the study workflow.

**Figure 1.**
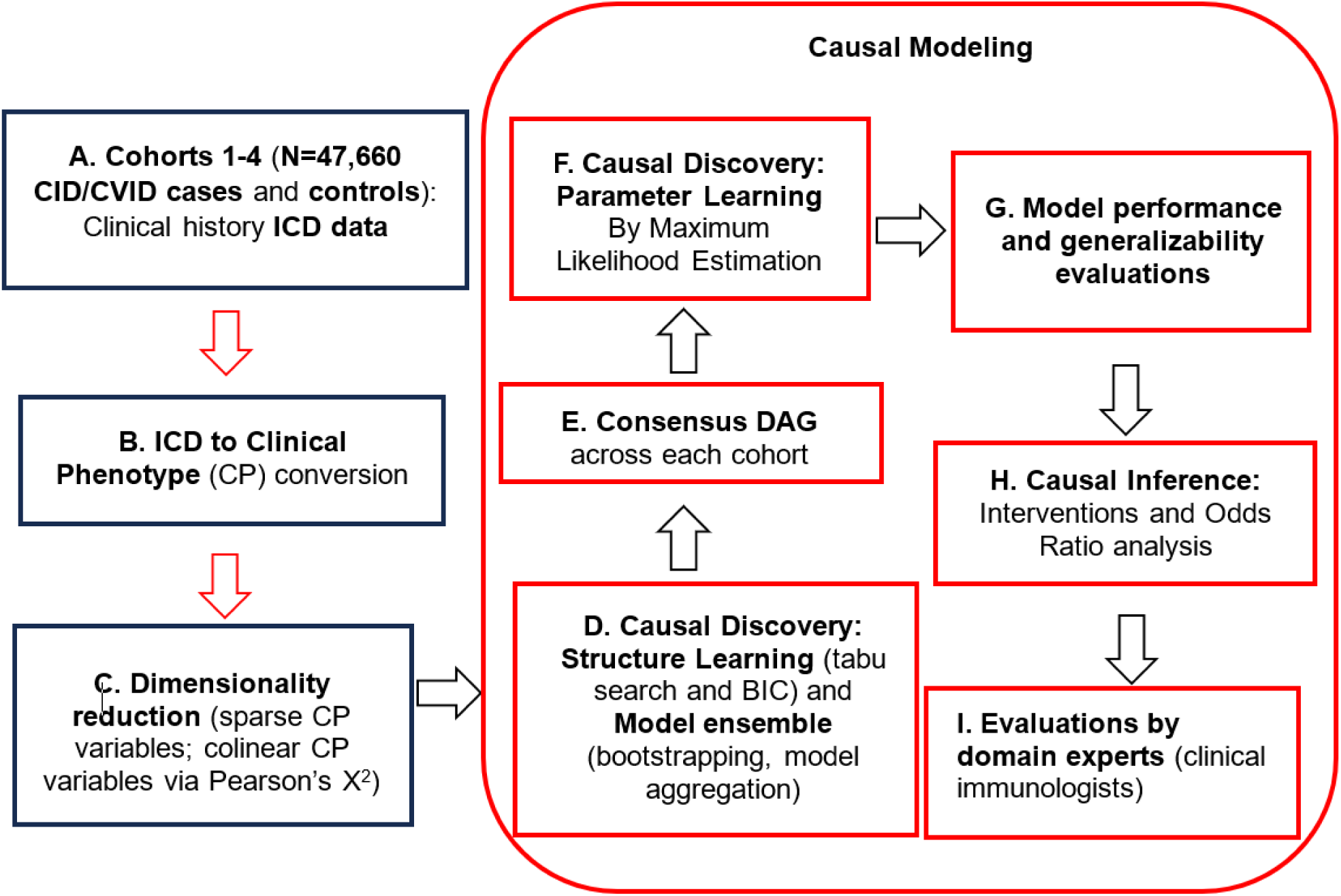
Study workflow. CID: Combined Immunodeficiency, CVID: Common Variable Immunodeficiency, BIC: Bayesian Information criterion, DAGs: directed acyclic graphs.

### Participants

Table 1 presents the patient demographics, which have been previously described [13]. In brief, age, gender, ethnicity, and patient history were similar between PI cases and controls. Most patients were female (53.1-62.2%) and Caucasian (82.4-88.1%). The mean age ranged from 44-48 years across cohorts. As anticipated, CID/CVID cases consistently had a higher number of healthcare visits compared to controls.

**Table 1.** Baseline demographics and clinical characteristics of the study participants (all four cohorts).

| Characteristics | Cohort 1 |  | Cohort 2 |  | Cohort 3 |  | Cohort 4 |  |
| --- | --- | --- | --- | --- | --- | --- | --- | --- |
|  | PI Cases<br>(N=797) | Controls<br>(N=797) | PI Cases<br>(N=797) | Controls<br>(N=797) | PI Cases<br>(N=2,312) | Controls<br>(N=2,312) | PI Cases<br>(N=19,924) | Controls<br>(N=19,924) |
| **Gender and Age** |  |  |  |  |  |  |  |  |
| Male (%) | 46.9 | 46.3 | 46.6 | 45.4 | 44.3 | 41.7 | 38.7 | 37.8 |
| Female (%) | 53.1 | 53.7 | 53.4 | 54.6 | 55.7 | 58.3 | 61.3 | 62.2 |
| Age (years) | 46 ± 26 | 46 ± 25 | 46 ± 26 | 48 ± 24 | 44 ± 26 | 45 ± 24 | 47 ± 24 | 46 ± 23 |
| 18-30 (%) | 16.39 | 16.32 | 16.98 | 17.02 | 17.12 | 17.14 | 13.89 | 14.01 |
| 31-50 (%) | 24.55 | 24.61 | 23.96 | 23.91 | 23.65 | 23.69 | 28.98 | 29.79 |
| 51-70 (%) | 35.85 | 35.79 | 36.05 | 35.85 | 34.81 | 34.79 | 35.76 | 36.03 |
| 71-max age (%) | 23.21 | 23.28 | 23.01 | 23.22 | 24.43 | 24.38 | 21.37 | 20.17 |
| **Patient History** |  |  |  |  |  |  |  |  |
| Diagnosis History duration (years) <sup>a</sup> | 10 (8-13) | 12 (10-14) | 10 (8-13) | 12 (9-14) | 9 (6-12) | 11 (8-14) | 9 (6-12) | 11 (8-14) |
| Number of visits <sup>a</sup> | 201 (103-399) | 145 (48-415) | 206 (105-399) | 182 (45-397) | 108 (36-250) | 73 (17-243.5) | 87 (30-206) | 64 (16-195) |
| **Ethnicity (%)** |  |  |  |  |  |  |  |  |
| African American | 8.7 | 7.3 | 8.4 | 6.7 | 7.9 | 7.2 | 5.8 | 5.8 |
| Asian | 1.4 | 1.5 | 1.3 | 0.5 | 1.7 | 1.6 | 1.2 | 1.1 |
| Caucasian | 82.4 | 85.1 | 83.4 | 88.1 | 81.7 | 84.2 | 85.3 | 86.8 |
| Other/Unknown | 7.5 | 6.1 | 7 | 4.7 | 8.7 | 7.1 | 7.8 | 6.3 |

<sup>a</sup> Median (25th–75th percentile)
PI: primary immunodeficiency (combined immunodeficiency in Cohorts 1-3; combined immunodeficiency and common variable immunodeficiency in Cohort 4).

BN models and their resulting consensus DAGs were generated in the setting of identifying CID/CVID patients against matched controls, across cohorts. All ICD codes were extracted from patient clinical histories and converted into clinical phenotypes, which were then used as inputs for the causal BN models. Initially, the model focused on identifying CID patients with pneumonia against matched controls (Cohort 1), then expanded to include controls without pneumonia (Cohort 2). Subsequently, the model was refined to identify all CID patients (Cohort 3) in our data and ultimately expanded to include all CID and CVID patients (Cohort 4), both against matched random controls. In Cohorts 3-4, cases and controls were selected irrespectively of pneumonia status. All controls were negative for CID, CVID, and PI.

### Consensus DAGs across cohorts

The cause-effect relationships in the consensus DAGs represent probabilistic associations, not strict chronological sequences: each parent phenotype significantly increases the likelihood of observing at least one of its child phenotypes in a patient’s history, regardless of their temporal order.

The consensus DAGs identified by performing causal discovery in Cohorts 1-4 are presented in Figures 2-5, respectively. Figures 2-5 illustrate: up to 2 direct parent levels and up to 2 direct child levels away from the CID/CVID diagnosis; and up to 1 direct parent level for each direct child and up to 1 direct child level for each direct parent. These consensus DAGs consistently reveal a network of interrelated comorbidities preceding CID/CVID diagnosis, including autoimmune and blood disorders, lymphomas, organ damage or inflammation, respiratory conditions, genetic anomalies, recurrent infections, and allergies.

**Figure 2.**
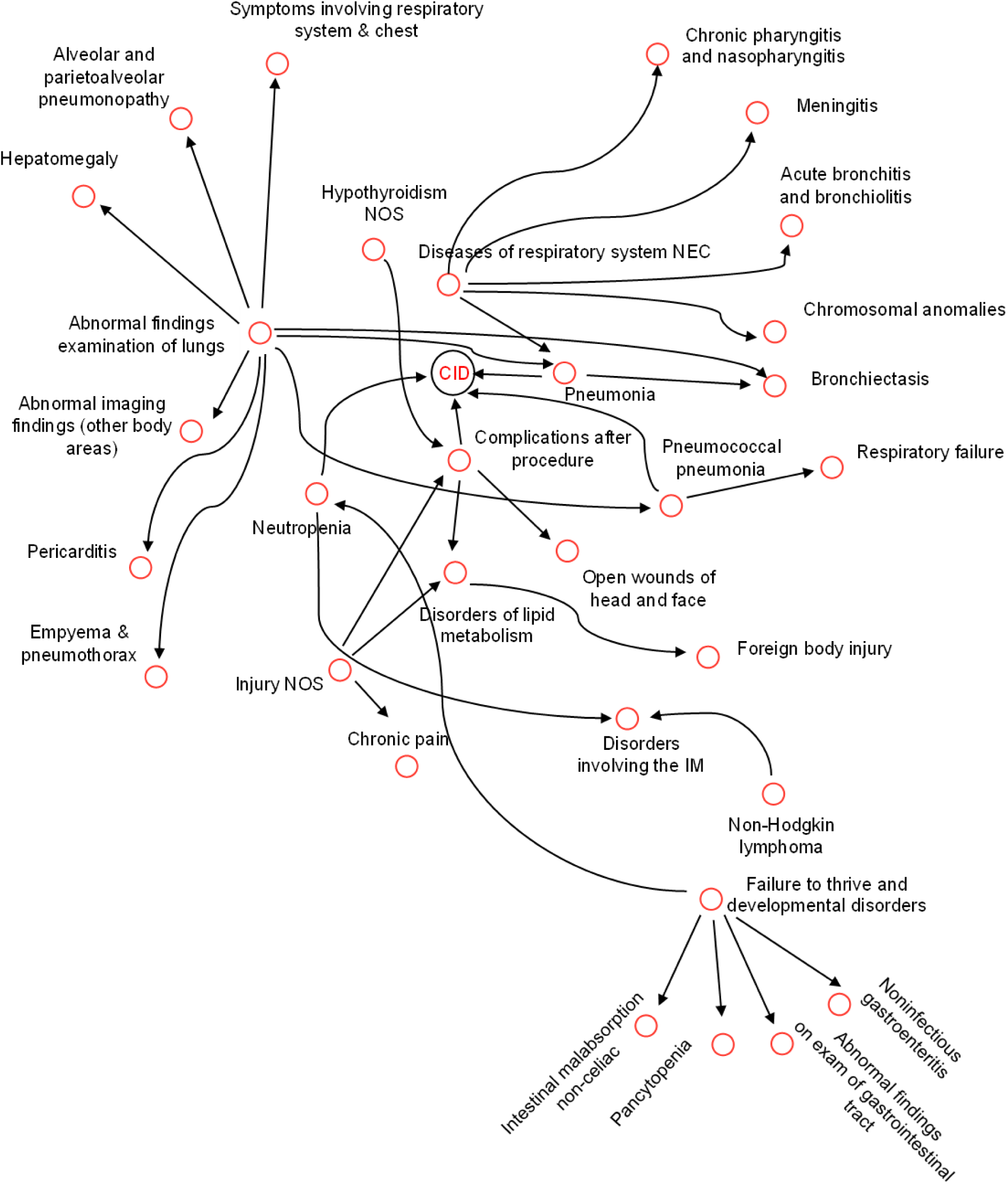
Consensus DAG calculated in Cohort 1. Cohort 1 involved N=797 CID cases with pneumonia and 797 matched controls (with no PI) with pneumonia. To improve clarity, we visualize up to 2 direct parent levels and up to 2 direct child levels away from CID diagnosis. To provide further context, up to 1 direct parent level for each direct child and up to 1 direct child level for each direct parent are included. DAG: directed acyclic graph; NEC: not elsewhere classified; NOS: not otherwise specified; IM: immune mechanism; CID: combined immunodeficiency; PI: primary immunodeficiency.

In Cohort 1, neutropenia, complications after procedure, pneumococcal pneumonia and general pneumonia were the direct parents of CID diagnosis (Figure 2). Abnormal findings from examinations on lungs and diseases of respiratory system not elsewhere classified (NEC) were the direct parents of multiple phenotypes including respiratory conditions (pneumococcal pneumonia, general pneumonia, bronchiectasis, empyema and pneumothorax, alveolar and parietoalveolar pneumonopathy, abnormal imaging findings, acute bronchitis and bronchiolitis), organ damage or inflammation (pericarditis, hepatomegaly) and infections or inflammations (meningitis, chronic pharyngitis and nasopharyngitis). Failure to thrive and developmental disorders was also the direct parent of gastrointestinal conditions and pancytopenia. Other phenotypes involved in this consensus DAG were non-Hodgkin lymphoma and disorders involving the immune mechanism.

In Cohort 2, neutropenia, bacterial pneumonia and influenza were the direct parents of CID diagnosis (Figure 3). Influenza, bacterial pneumonia, abnormal findings from examinations on lungs and acute pharyngitis were the direct parents of multiple phenotypes including respiratory conditions (bronchopneumonia and lung abscess, pseudomonal pneumonia, empyema and pneumothorax, acute bronchitis and bronchiolitis, pulmonary inflammation or edema, bronchiectasis, pneumococcal pneumonia), acute or recurrent infections (acute sinusitis, chronic tonsilitis and adenoiditis, acute pharyngitis, otitis media, skin infections, bacteremia, meningitis, candidiasis, mycoses), allergies or allergic reactions (allergic rhinitis, urticaria), organ inflammation (pericarditis), non-Hodgkin lymphoma, developmental delays/ disorders and disorders of the immune system (IM; the latter typically associated with autoimmune diseases [13]).

**Figure 3.**
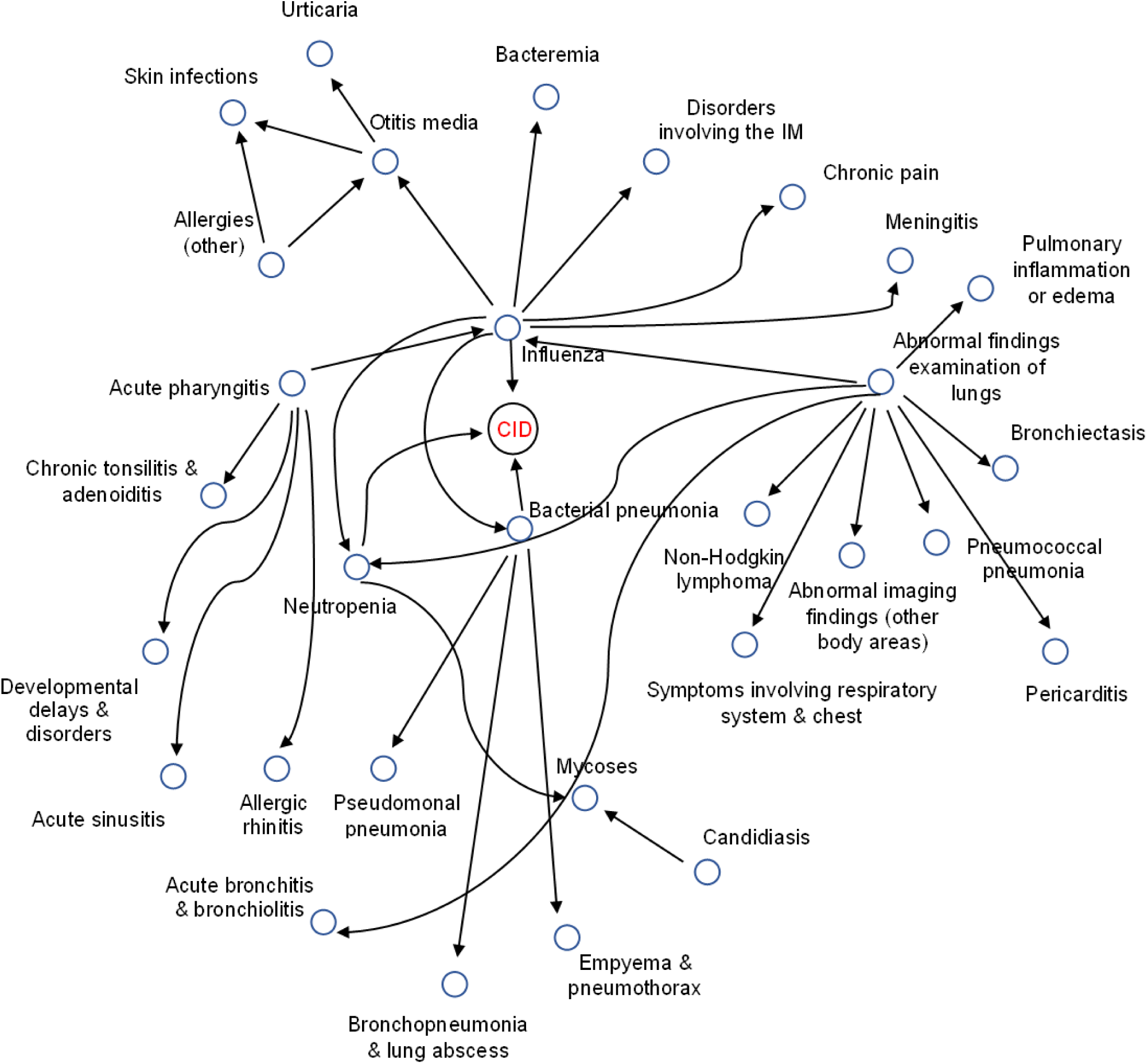
Consensus DAG calculated in Cohort 2. Cohort 2 involved N=797 CID cases with pneumonia and 797 matched controls (with no PI) with and without pneumonia. We visualize up to 2 direct parent levels and up to 2 direct child levels away from CID diagnosis. Up to 1 direct parent level for each direct child and up to 1 direct child level for each direct parent are included. DAG: directed acyclic graph; IM: immune mechanism; CID: combined immunodeficiency; PI: primary immunodeficiency.

In Cohort 3, neutropenia, genetic susceptibility to disease and encounter for long-term use of antibiotics were the direct parents of CID (Figure 4). Developmental delays/ disorders, abnormal findings from examinations on lungs, bacterial infection not otherwise specified (NOS), disorders involving the immune mechanism and acute bronchitis and bronchiolitis were the direct parents of several phenotypes including acute or chronic respiratory conditions (pleurisy and pleural effusion, respiratory failure, emphysema), infections (sepsis, bacteremia, acute sinusitis, acute pharyngitis, streptococcus infection), gastrointestinal conditions (gastritis and duodenitis, diarrhea), blood conditions (decreased white blood cell count (bcc), anemia of chronic disease) and organ damage (splenomegaly). Other phenotypes identified in the consensus DAG were non-Hodgkin lymphoma, symptoms concerning nutrition, metabolism and development, failure to thrive and edema.

**Figure 4.**
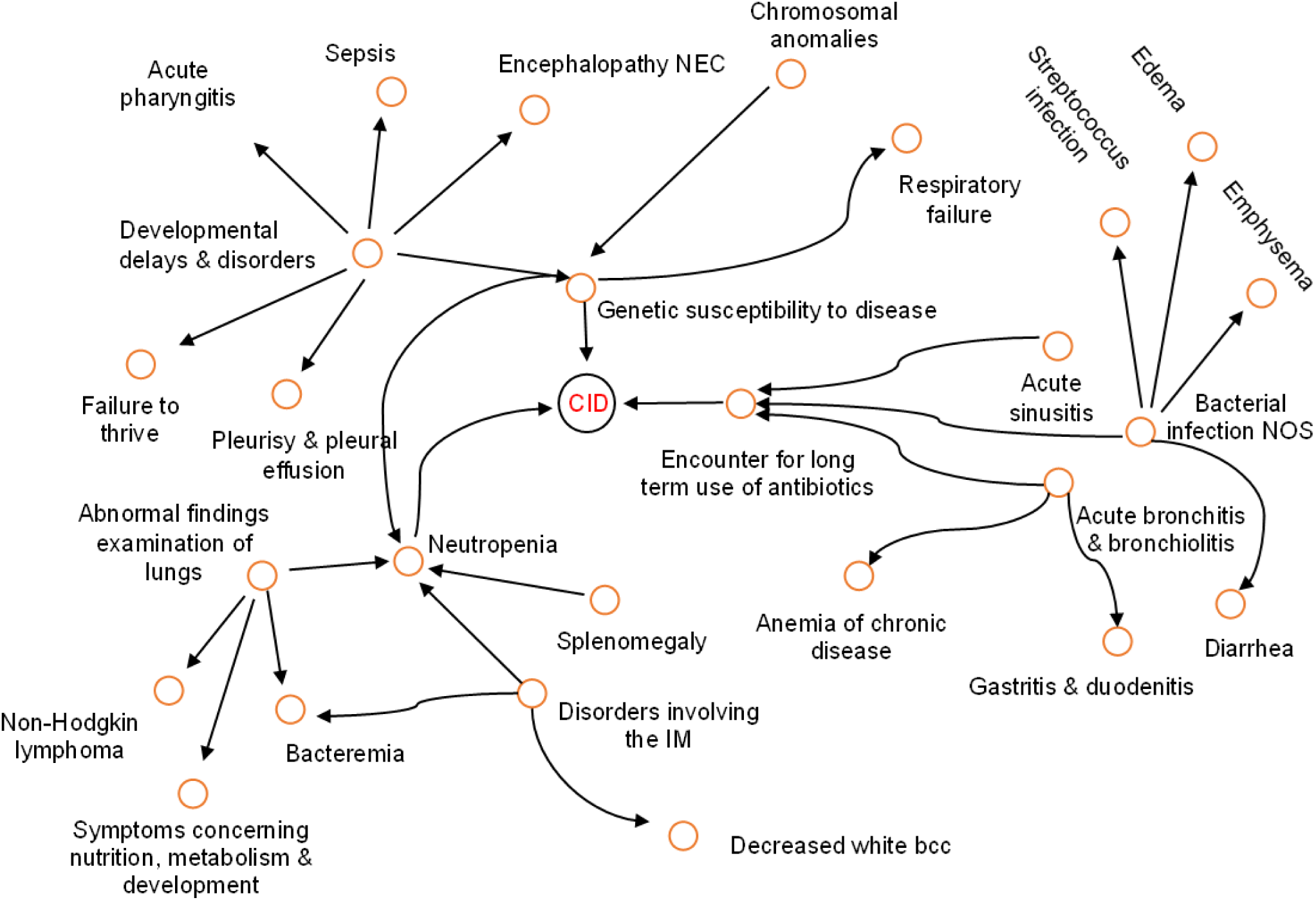
Consensus DAG calculated in Cohort 3. Cohort 3 involved N=2,312 CID cases (of which 797 with pneumonia) and 2,312 matched controls (with no PI), both with and without pneumonia. We visualize up to 2 direct parent levels and up to 2 direct child levels away from CID diagnosis. Up to 1 direct parent level for each direct child and up to 1 direct child level for each direct parent are included. DAG: directed acyclic graph; NEC: not elsewhere classified; NOS: not otherwise specified; IM: immune mechanism; bcc: blood cell count; CID: combined immunodeficiency; PI: primary immunodeficiency.

In Cohort 4, autoimmune disease NEC, hypothyroidism NOS, neutropenia, developmental delays/ disorders and bronchiectasis were the direct parents of CID/CVID diagnosis (Figure 5). In turn, hypothyroidism NOS, bronchiectasis, neutropenia, viral infection and bacterial pneumonia were the direct parents of multiple phenotypes including acute or chronic respiratory conditions (bronchitis, asthma, asphyxia and hypoxemia), infections or inflammations (chronic sinusitis, bacteremia, otitis media, chronic pharyngitis and nasopharyngitis) autoimmune diseases (rheumatoid arthritis), gastrointestinal conditions (gastritis and duodenitis, non-infectious gastroenteritis) and allergies (allergic rhinitis). Other phenotypes involved were non-Hodgkin lymphoma and abnormal electrocardiogram (ECG).

**Figure 5.**
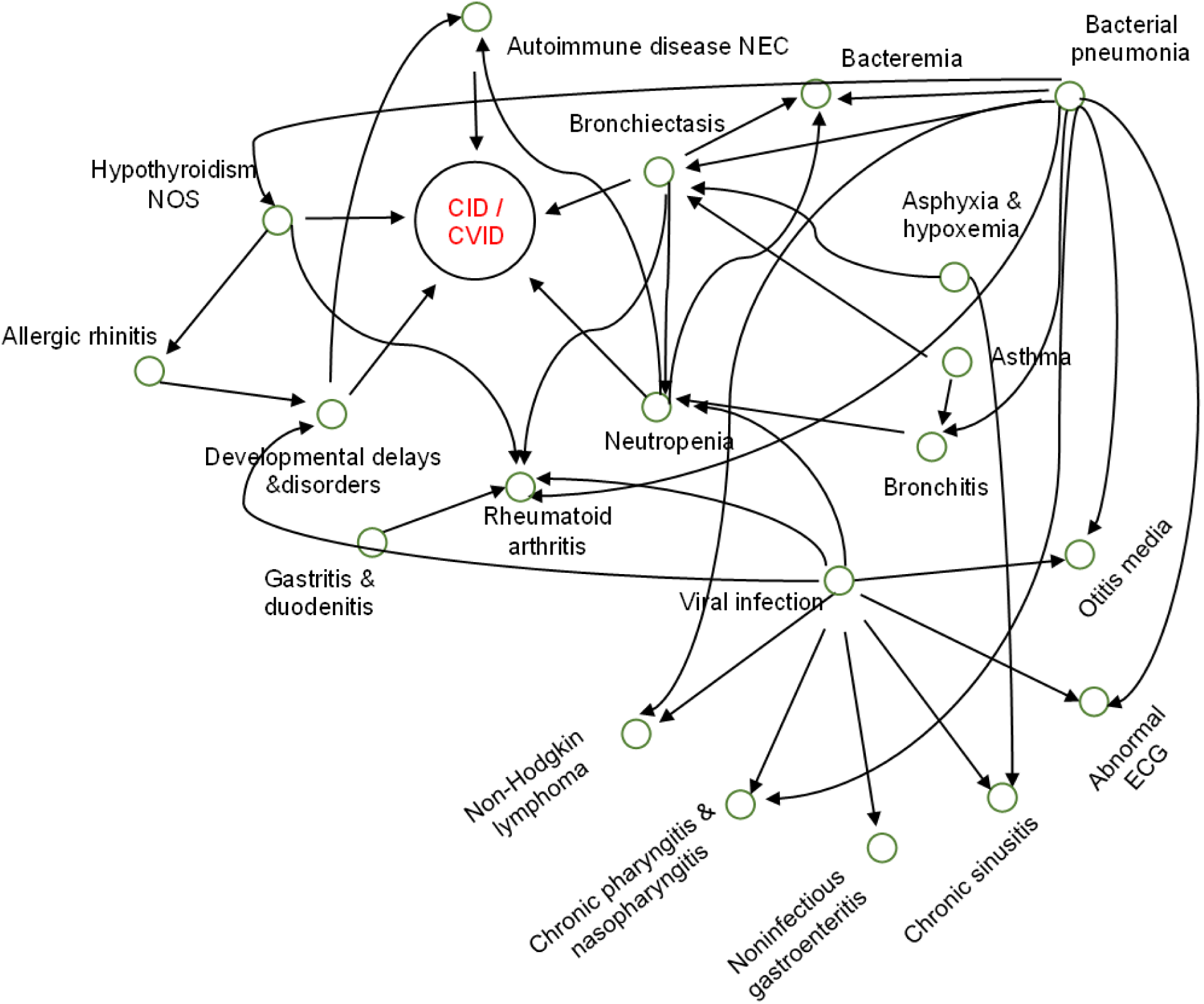
Consensus DAG calculated in Cohort 4. Cohort 4 involved N=19,924 CID and CVID cases (of which 2,350 with pneumonia) and 19,924 matched controls (with no PI), both with and without pneumonia. We visualize up to 2 direct parent levels and up to 2 direct child levels away from CID diagnosis. Up to 1 direct parent level for each direct child and up to 1 direct child level for each direct parent are included. DAG: directed acyclic graph; NEC: not elsewhere classified; NOS: not otherwise specified; IM: immune mechanism; ECG: electrocardiogram; CID: combined immunodeficiency; CVID: common variable immunodeficiency; PI: primary immunodeficiency.

### Predictive accuracy within the same population

Subsequently, we evaluated the predictive ability of each consensus DAG in identifying CID/CVID in an unseen test set from the same population. ROC analysis showed good predictive performance within each cohort (Table 2, Figure 6).

**Figure 6.**
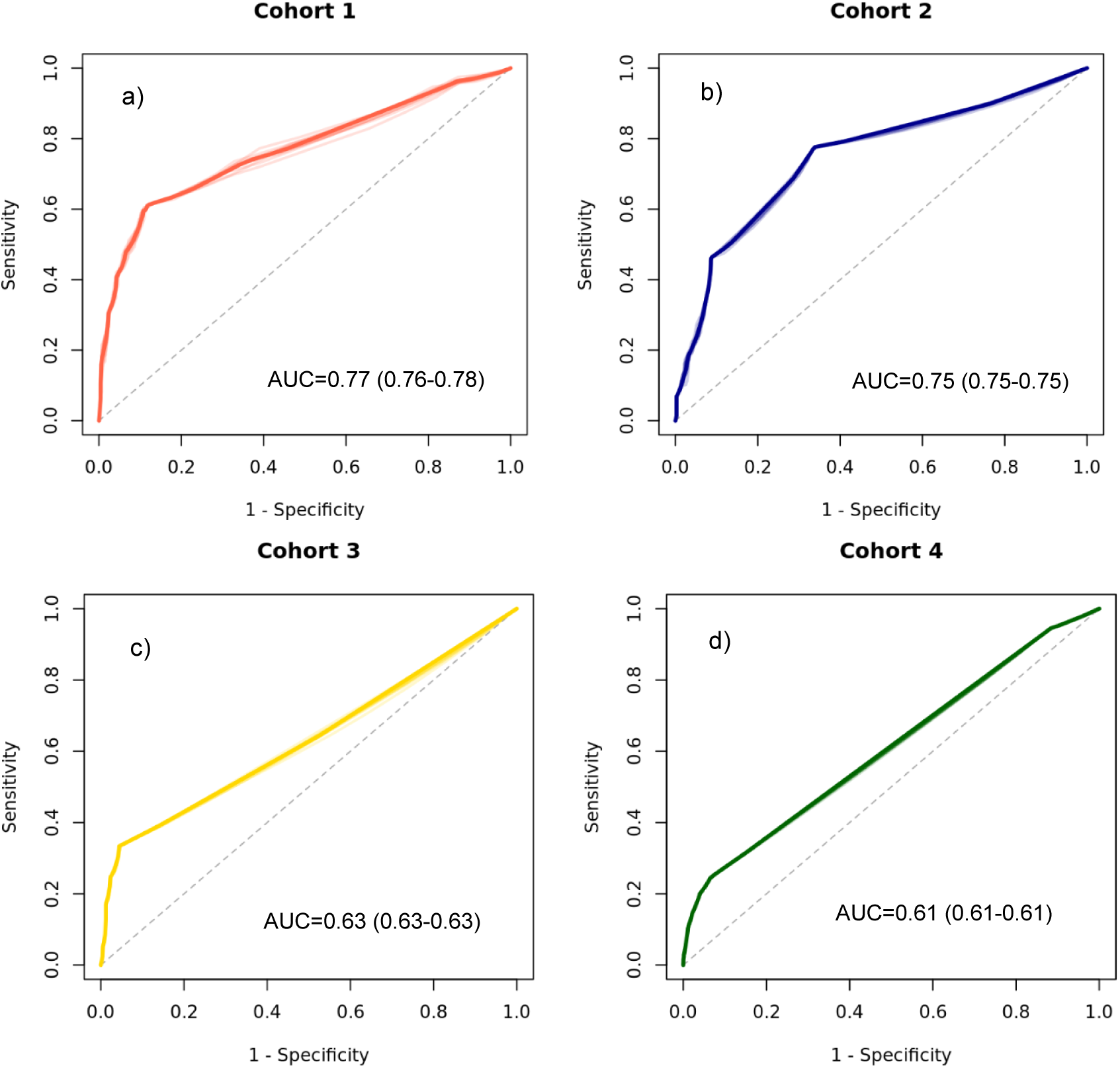
Receiver operating characteristic curves (ROC) for all causal models developed (in the training set) and evaluated (test set) across all four cohorts. Here, ROC analysis demonstrates the evaluations performed in the held-out test set, within each cohort (e.g., a DAG trained and tested in Cohort 1, a DAG trained and tested in Cohort 2, and so forth). a) CID patients with pneumonia against pneumonia patients without PI (N = 1594; 797 CID cases and 797 controls). b) CID patients with pneumonia against randomly selected patients without PI, with and without pneumonia (N = 1594; 797 CID cases and 797 controls). c) CID patients with and without pneumonia against randomly selected patients without PI, with and without pneumonia (N = 4624; 2312 CID cases and 2,312 controls). d) All CID and CVID patients with and without pneumonia against randomly selected patients without PI, with and without pneumonia (N = 39,848; 19,924 PI cases and 19,924 controls). Across all cohorts, PI cases and controls were 1:1 matched for age, gender, race, ethnicity, duration of medical history, and the number of healthcare visits. CID: combined immunodeficiency; CVID: common variable immunodeficiency.

**Table 2.** Mean diagnostic performance of causal modeling predictions in the testing set, across all evaluations performed in Cohorts 1-4. Within cohort evaluations in the held-out fold test set are shown with bold. Parentheses show standard deviation.

| Patient Cohorts |  |  |  |  |  |
| --- | --- | --- | --- | --- | --- |
| Metric Trained on (column)/<br>Predicted on (row) |  | Cohort 1 | Cohort 2 | Cohort 3 | Cohort 4 |
| Sensitivity | Cohort 1 | **0.84 (0.00)** | 0.69 (0.01) | 0.78 (0.00) | 0.79 (0.00) |
|  | Cohort 2 | 0.83 (0.00) | **0.70 (0.00)** | 0.81 (0.00) | 0.78 (0.00) |
|  | Cohort 3 | 0.74 (0.00) | 0.67 (0.00) | **0.88 (0.00)** | 0.76 (0.00) |
|  | Cohort 4 | 0.76 (0.00) | 0.66 (0.01) | 0.69 (0.00) | **0.78 (0.00)** |
| Specificity | Cohort 1 | **0.69 (0.00)** | 0.63 (0.00) | 0.62 (0.00) | 0.61 (0.00) |
|  | Cohort 2 | 0.67 (0.00) | **0.75 (0.01)** | 0.63 (0.01) | 0.61 (0.00) |
|  | Cohort 3 | 0.55 (0.01) | 0.55 (0.01) | **0.59 (0.00)** | 0.56 (0.00) |
|  | Cohort 4 | 0.54 (0.01) | 0.54 (0.01) | 0.55 (0.00) | **0.55 (0.00)** |
| Accuracy | Cohort 1 | **0.75 (0.00)** | 0.65 (0.00) | 0.67 (0.00) | 0.66 (0.01) |
|  | Cohort 2 | 0.73 (0.00) | **0.72 (0.00)** | 0.69 (0.00) | 0.66 (0.00) |
|  | Cohort 3 | 0.58 (0.01) | 0.57 (0.00) | **0.65 (0.00)** | 0.59 (0.01) |
|  | Cohort 4 | 0.57 (0.01) | 0.56 (0.01) | 0.58 (0.00) | **0.59 (0.00)** |
| ROC AUC | Cohort 1 | **0.77 (0.01)** | 0.66 (0.00) | 0.71 (0.00) | 0.65 (0.00) |
|  | Cohort 2 | 0.72 (0.00) | **0.75 (0.00)** | 0.72 (0.01) | 0.65 (0.00) |
|  | Cohort 3 | 0.58 (0.01) | 0.59 (0.00) | **0.63 (0.00)** | 0.56 (0.00) |
|  | Cohort 4 | 0.57 (0.00) | 0.56 (0.01) | 0.59 (0.00) | **0.61 (0.00)** |

In Cohorts 1-2, the model achieved strong predictive performance with a sensitivity of 0.84 and 0.70, a specificity of 0.69 and 0.75, overall accuracy of 0.75 and 0.72 and an AUC of 0.77 and 0.75, respectively. In Cohorts 3-4, the model showed good predictive performance with a sensitivity of 0.88 and 0.78, a specificity of 0.59 and 0.55, overall accuracy of 0.65 and 0.59 and an AUC of 0.63 and 0.61, respectively.

### Generalizability to other populations

When the consensus DAG models were applied to unseen data from other cohorts, they maintained high predictive accuracy across all evaluations (Table 2). Sensitivity, specificity, accuracy, and AUC ranged from 0.83-0.66, 0.67-0.54, 0.73-0.57, and 0.72-0.56 respectively.

Notably, the consensus DAG models trained on larger cohorts (Cohorts 3-4) showed improved predictive performance when tested in unseen smaller data (Cohorts 1-2), against when tested in unseen data from the cohorts they were trained on (Table 2). Conversely, models trained on smaller cohorts (Cohorts 1-2) demonstrated reduced predictive performance when applied to new (larger) data.

### Causal inference

Interventional analysis identified key antecedent phenotypes with high ORs (Table 3). The following antecedent phenotypes with ORs greater than 2.00 were identified in each cohort: Cohort 1: pneumococcal pneumonia, neutropenia and general pneumonia (OR range: 13.09-4.09); Cohort 2: neutropenia, bacterial pneumonia and influenza (OR range: 6.07-3.55); Cohort 3: failure to thrive, genetic susceptibility to disease, disorders involving the IM and decreased white bcc (OR range: 23.65-5.14). Cohort 4; bronchiectasis, autoimmune disease NEC, neutropenia and developmental delays/ disorders (OR range: 9.44-2.25).

**Table 3.** Temporal information and interventions (causal inference) in Cohorts 1 and 2. Temporal distributions were calculated by considering the first diagnosis of each phenotype in reference to the first CID diagnosis, in terms of Box and Whisker plots: median value and 50% interquartile range to the median (lower and higher interquartile value to the median, shown in parenthesis). All temporal information (median values and interquartile ranges) is expressed in months. Odds ratio represents the effect of each intervention on the CID diagnosis. NEC: not elsewhere classified; NOS: not otherwise specified.

| Phenotype per cohort | Temporal Information<br>(in months) | Intervention<br>(Odds ratio) |
| --- | --- | --- |
| **Cohort 1** |  |  |
| Pneumococcal pneumonia | -0.13 (-11.4, -10.61) | 13.09 |
| Neutropenia | -10.3 (-36.0, 0.2) | 7.22 |
| Pneumonia | -14.3 (-46.7, 0.0) | 4.09 |
| Abnormal findings examination of lungs | -10.0 (-32.8, 0.2) | 1.67 |
| Failure to thrive and developmental disorders | -14.5 (-48.4, 0.0) | 1.29 |
| Diseases of respiratory system NEC | -14.9 (-45.0, 1.5) | 1.29 |
| Bacteremia | -1.7 (-29.9, 5.3) | 1.11 |
| Pancytopenia | -10.6 (-34.4, 0.4) | 1.02 |
| Non-Hodgkin lymphoma | -21.9 (-60.3, -0.4) | 1.02 |
| Meningitis | -34.1 (-61.1, -11.8) | 1.02 |
| **Cohort 2** |  |  |
| Neutropenia | -10.3 (-35.4, 0.3) | 6.07 |
| Bacterial pneumonia | -1.6 (-22.9, 3.1) | 6.06 |
| Influenza | -7.9 (-30.3, 14.15) | 3.55 |
| Abnormal findings examination of lungs | -9.7 (-32.7, 0.6) | 1.70 |
| Acute pharyngitis | -19.2 (-55.9, 5.5) | 1.33 |
| Bacterial infection NOS | -3.1 (-26.5, 8.2) | 1.19 |
| Viral infection | -17.2 (-50.7, 2.6) | 1.04 |
| Allergies | -14.0 (-41.5, 5.4) | 1.02 |
| Acute bronchitis | -19.5 (-45.6, 0.0) | 1.02 |
| Otitis media | -15.9 (-36.7, 9.3) | 1.01 |

### Qualitative evaluation by clinical immunologists

Three clinical immunologists (RT, VHT, JR) reviewed the consensus DAG outcomes (Figures 2-5, Tables 2-3) against their clinical experience and prior studies on PI [13, 14, 39, 40]. All 3 clinicians agreed that the DAGs could substantially enhance patient screening by identifying phenotype combinations on the following trajectories:

a. Direct precursors of CID/CVID diagnoses (e.g., bacterial pneumonia) alongside conditions from different phenotype families (e.g., acute pharyngitis) or disease complications (e.g., bronchiectasis), anywhere in the DAG.
b. Parent phenotypes (e.g., abnormal findings in examinations of lungs) associated with child phenotypes from different phenotype families (e.g., pericarditis, hepatomegaly, lymphoma, meningitis) or disease complications (e.g., sepsis).
c. Associations between parent phenotypes and other, not necessarily interconnected, child phenotypes from different phenotype families or disease complications.

The consensus among clinicians was that certain phenotypes identified in (a-c) may be subject to recurrence, aligning with existing medical literature on the recurrent nature of conditions such as pneumonias, infections, and inflammations [1–10, 13].

According to all clinicians, analysis of consensus DAGs in the context of prior large-scale studies [13, 14, 39, 40] revealed a broader and more nuanced spectrum of PI-associated comorbidities that could precede CID/CVID diagnosis, potentially enhancing their identification.

## Discussion

In this study, we present a novel approach to identify antecedent patient comorbidities associated with CID and CVID, through the development and evaluation of consensus DAGs derived from BN models. Our findings demonstrate that these DAGs can effectively identify CID/CVID diagnoses across diverse patient cohorts, exhibiting good predictive accuracy both within the training population and when generalized to unseen populations. Notably, this methodology offers a unique advantage by revealing complex interrelationships among a wide array of comorbidities preceding CID/CVID diagnosis, including autoimmune and blood disorders, lymphomas, organ damage or inflammation, respiratory conditions, genetic anomalies, recurrent infections and allergies. This comprehensive understanding of the antecedent phenotypic landscape has the potential to significantly improve patient screening and early detection of these PIs.

To the best of our knowledge, this is the first study to apply causal discovery methods to clinical history phenotypes derived from diagnosis codes, and the first such investigation within the context of PI. While not directly pertinent to causal discovery, a previous study employed a BN structure to quantify the interplay of diagnosis codes within a pediatric cohort (N=3,460 patients and 1:1 matched controls) [18]. However, this approach relied on a predetermined set of 36 diagnosis codes selected by an expert immunologist, potentially introducing bias into the BN structure (due to involving a single domain expert) and limiting its generalizability to larger and more clinically diverse patient populations. Prior research has demonstrated the efficacy of ML models in identifying PI, including CID and CVID, using EHR-derived clinical history diagnosis codes [13, 14]. In our recent work, we demonstrated that ML models can identify CID and CVID with high accuracy [13], from the same populations used in our current work. By using descriptive statistics, we have also identified combinations of antecedent phenotypes associated with these conditions [13]. Building upon our prior work [13], but without imposing any knowledge from it, our causal discovery method has independently identified, represented and interrelated many of these antecedent phenotypes within the consensus DAGs across cohorts (Figures 2-5, Tables 3-4). Among these, our interventional analysis identified key antecedent phenotypes (respiratory conditions, blood disorders, developmental delays, autoimmune diseases) with high ORs (Table 3). Our causal discovery methodology can offer a distinct advantage by explicitly unveiling probabilistic trajectories across clinical history phenotypes, which can be used to potentially improve the early suspicion and identification of adult patients at risk for CID/CVID.

**Table 4.** Temporal information and interventions (causal inference) in Cohorts 3 and 4. Temporal distributions were calculated by considering the first diagnosis of each phenotype in reference to the first CID/CVID diagnosis, in terms of Box and Whisker plots: median value and 50% interquartile range to the median (lower and higher interquartile value to the median, shown in parenthesis). All temporal information (median values and interquartile ranges) is expressed in months. Odds ratio represents the effect of each intervention on the CID/CVID diagnosis. IM: immune mechanism; bcc: blood cell count; NEC: not elsewhere classified.

| Phenotype per cohort | Temporal Information<br>(in months) | Intervention<br>(Odds ratio) |
| --- | --- | --- |
| **Cohort 3** |  |  |
| Failure to thrive | -4.9 (-31.0, 0.7) | 23.65 |
| Genetic susceptibility to disease | -4.1 (-30.7, 1.2) | 13.24 |
| Disorders involving the IM | -8.2 (-35.2, -0.4) | 8.74 |
| Decreased white bcc | -3.03 (-25.4, 0.7) | 5.14 |
| Splenomegaly | -4.9 (-24.7, 1.8) | 1.49 |
| Nutritional, metabolic, and developmental symptoms | -10.0 (-34.9, 0.9) | 1.46 |
| Developmental delays & disorders | -5.8 (-33.0, 6.3) | 1.26 |
| Pancytopenia | -5.2 (-29.5, 0.7) | 1.22 |
| Pneumonia | -13.7 (-46.6, 0.0) | 1.16 |
| Autoimmune disease NEC | -28.8 (-61.2, -2.2) | 1.16 |
| **Cohort 4** |  |  |
| Bronchiectasis | -4.2 (-29.6, 1.3) | 9.44 |
| Autoimmune disease NEC | -23.7 (-50.1, -3.2) | 4.36 |
| Neutropenia | -4.9 (-29.8, 0.9) | 3.68 |
| Developmental delays & disorders | -7.8 (-39.3, 5.7) | 2.25 |
| Bacterial pneumonia | -1.8 (-23.4, 5.8) | 1.14 |
| Asphyxia and hypoxemia | -2.1 (-25.3, 6.5) | 1.13 |
| Asthma | -27.9 (-62.8, -4.5) | 1.10 |
| Bronchitis | -7.8 (-19.8, -0.2) | 1.08 |
| Viral infection | -16.5 (-45.8, 0.3) | 1.07 |
| Rheumatoid arthritis | -17.2 (-61.8, -0.2) | 1.01 |

Standard (non-causal) machine learning (ML) and statistical models frequently fail to capture the intricate interplay and probabilistic dependencies among variables (phenotypes), thereby potentially limiting their generalizability, robustness, and interpretability [15–17, 41]. Of note, previous ML research on large-scale PI datasets consisting of patient clinical history (diagnosis codes), has primarily focused on evaluating the predictive performance of ML models on unseen data drawn from the same population used for model training [13, 14]. Regarding generalizability, without the capacity to discern causal mechanisms and spurious associations, the predictive accuracy of non-causal ML and statistical models is compromised when the distribution of the testing data diverges from that of the training data [15, 16]. It is known that variations in the sampled populations, such as the patient characteristics and clinical histories observed in Cohorts 1-4, can potentially degrade model generalizability if the model was not exposed to such variations during development [15–17, 41–43]. This issue, referred to as the out-of-distribution generalization challenge in ML, constitutes an active research area, with causal modeling identified as a potential solution to mitigate these limitations [15]. Our results support these methodological developments, demonstrating the robust performance of consensus DAG models across diverse cohorts, including those not represented in the training data (Table 2). While maintaining high predictive accuracy within the same cohort, the models exhibited notable generalizability across datasets. Importantly, consensus DAG models trained on the largest and most heterogeneous cohorts (Cohorts 3-4) showed superior performance on smaller, unseen datasets (Cohorts 1-2) compared to their performance on unseen data from the cohorts they were originally trained on (Table 2). By giving access to our open-source code, learning consensus DAGs across further large external CID/CVID cohorts, and potentially other PI subtypes, could provide important clinical utility by enabling the generation of informative consensus DAGs in the setting of predicting PI in smaller cohorts e.g., derived from certain patient populations or healthcare systems. Moreover, our analysis indicates that causal modeling, by accounting for underlying causal mechanisms across phenotype occurrences, enhances model robustness and generalizability across diverse data distributions, thereby addressing the out-of-distribution generalization challenge in our data. By incorporating causal discovery into our screening and early detection tool, we can potentially enhance its generalizability, robustness, and interpretability, ultimately contributing to more effective clinical decision-making and improved patient outcomes.

In terms of clinical interpretation of the constructed consensus DAGs, the presence of a parent phenotype in a patient’s clinical history signals a heightened likelihood of observing its child phenotypes, regardless of their chronological order. This interpretation suggests that the DAG can be utilized as a diagnostic tool for identifying groups of patients who may exhibit specific clusters of phenotypes, even if these phenotypes do not appear in a strict temporal sequence. Consequently, the DAG could serve as a valuable resource for clinicians, potentially aiding in early suspicion and diagnosis of CID/CVID, by highlighting key phenotypic trajectories within patient histories.

Our study elucidates a consistent pattern of interconnected comorbidities preceding CID/CVID diagnosis, demonstrating a complex interplay of factors contributing to their clinical manifestation. While the specific phenotypes directly preceding CID/CVID diagnosis varied across cohorts (reflecting differences in patient populations), the broader constellations of antecedent conditions remained remarkably consistent. This highlights the robustness of our causal discovery approach and suggests the presence of shared underlying causal history trajectories across diverse patient populations. Notably, neutropenia emerges as a key antecedent and direct parent of CID/CVID across all cohorts, suggesting that it may be an early clinical indicator or risk factor for these conditions. This aligns with existing literature highlighting the association between neutropenia and PI, further underscoring its clinical relevance [40]. The prominent involvement of respiratory conditions and complications, infections, and inflammatory processes across cohorts aligns with the known susceptibility of individuals with PIs to these manifestations, reinforcing the importance of early PI identification and management [1–10, 12]. The presence of allergies across multiple cohorts is consistent with the known association between allergic manifestations and PI [1–5, 10, 44]. Additionally, the presence of multiple autoimmune diseases highlights a known link between autoimmunity and PIs [1–5, 10, 12, 40, 44]. Ongoing research aims to elucidate the precise genetic and immunological mechanisms underpinning the relationships between autoimmunity and PIs [45]. The identification of developmental disorders as precursors across all cohorts is in line with current medical knowledge of inherited or early-life factors in individuals with CID/CVID [1–5]. Furthermore, the presence of non-Hodgkin lymphoma in all cohorts highlights the established link between PI and increased risk for lymphoid malignancies [1–5, 12, 40], emphasizing the need for heightened surveillance in this patient population to address both such severe co-morbidities and PI. The consistent identification of gastrointestinal disorders across cohorts aligns with the established link between PIs and such antecedent manifestations [1–3, 40, 44]. These findings provide a comprehensive, data-driven understanding of the complex network of comorbidities associated with CID/CVID, offering valuable insights for early detection, risk stratification, and personalized treatment strategies. The consistent patterns identified across cohorts further emphasize the potential of causal discovery methods to uncover meaningful relationships within clinical data and inform clinical practice.

Several limitations warrant consideration in the interpretation of our findings. The major limitation lies in the reliance on a set of assumptions necessary for conducting causal modeling (described in our Methods). Specifically, observational studies such as ours face the inherent limitation of partial identifiability [29, 30]. This can result in ambiguity in causal direction, as multiple causal models may fit the observed data equally well. In addition, the critical assumptions of faithfulness and the absence of unobserved variables, while theoretically necessary for interpreting arcs as causal effects, cannot be statistically verified [16]. Violations of these assumptions can lead to misinterpretations of causal relationships. However, to mitigate these limitations and ensure a robust interpretation of our findings, we employed a multi-faceted approach which included: a) accurate predictive performance on held-out test data within and across cohorts created by using different inclusion criteria, to assess model performance and generalizability, respectively; b) incorporation of expert knowledge from clinical immunologists to enhance the validity of causal interpretations; c) causal inference through interventions on BN variables to observe their effects on the odds of CID/CVID diagnosis, providing further empirical support for our causal claims; d) an ensemble approach to reduce bias and variance across individual DAGs, ultimately identifying the most prevalent variables within the consensus DAGs [16, 36].

In conclusion, our study demonstrates the potential of causal BNs to uncover complex trajectories among clinical phenotypes preceding CID/CVID diagnosis. The consensus DAGs exhibit robust predictive performance and generalizability across diverse patient cohorts, offering a promising avenue for enhanced screening and early detection of these conditions. Our multi-pronged approach, incorporating BN model predictions across diverse cohorts, causal inference and expert knowledge, strengthens the validity and clinical relevance of our findings. The identified phenotypic trajectories and their causal relationships hold considerable promise for translating into improved clinical practice, potentially leading to earlier identification and intervention for adults at risk of CID/CVID.

## Methods

### Dataset extraction and curation

To characterize patient history and perform causal discovery, we used International Classification of Diseases (ICD) diagnosis codes (medical claims) extracted from large anonymized Electronic Health Records (EHR) (Optum®, Inc., Eden Prairie, MN), a US nationally representative cohort covering all 50 U.S. States. The study was performed with the approval of Pfizer US Medical Affairs Hospital Specialty Care Leadership Team. Data extraction, pre-processing, causal modeling and evaluation of the Optum data were performed in accordance with the Declaration of Helsinki. The Optum data have been acquired according to the Health Insurance Portability and Accountability Act (HIPAA) Privacy Rule and all data were fully de-identified before licensed by Pfizer [13].

The end-to-end process of ICD data extraction and curation has been previously described [13]. Our ICD data spanned from January 1, 2008 to December 31, 2021 and included diagnosis codes from approximately 100 million US patients, featuring detailed records of clinical histories and demographic information. Clinical history ICD codes were converted into clinical phenotypes which were then utilized as data inputs for causal discovery modeling (see details in the subsection “Converting ICD codes to phenotypes”). Demographic information was used to match cases and controls using propensity score matching [13]. Participants were divided into four distinct cohorts (Cohorts 1-4), consisting of 797, 797, 2,312 and 19,924 PI cases respectively, with each cohort having an equivalent number of controls (a total of N= 47,660 cases and controls). The inclusion criteria required participants to be at least 18-years old at the time of PI diagnosis [13].

The identification of CID and CVID was based on ICD codes obtained from https://www.icd10data.com/, by including all D81 (for CID) and D83 (CVID) sections and subsections [13]. Supplementary Table 1 details all the ICD codes for CID/CVID, as identified in the Optum database at the time of our data extraction.

In all cohorts, cases of PI and controls were 1:1 matched for age, gender, race, ethnicity, duration of medical history (in months) and number of healthcare visits, through propensity score matching. This resulted in an even distribution of PI patients and PS-matched controls within each cohort. Across each patient and control in Cohorts 1-4, all available ICD codes were extracted and added in the list of clinical history [13]. The presence or absence of all ICD codes identified were used as binary categorical features.

### Cohort generation

As previously described [13], given that pneumonia is the most frequent severe infection in CID [1, 8–10], we first generated BN models to identify CID patients with pneumonia against matched controls with pneumonia (Cohort 1). We then generated another set of BN models to identify CID patients with pneumonia against matched controls with or without pneumonia (Cohort 2). We continued BN model development by aiming to identify CID patients against matched random controls (both with or without pneumonia) (Cohort 3). Lastly, we expanded our dataset and developed another set of BN models to identify both CID and CVID patients against matched random controls (both with or without pneumonia). Across all cohorts, we ensured none of the controls had CID, CVID or PI.

### ICD data preparation

Across all Cohorts 1-4, ICD-10 / ICD-9 codes and patient demographics were mined from the Optum® patient and diagnosis tables using Dataiku: https://www.dataiku.com/ [13]. All the ICD-9 codes present in the data were converted to ICD-10, using the updated general equivalence mappings (2018 GEMS) from the https://www.cms.gov/ website, as previously described [13]. All ICD-10 codes were then converted to disease descriptions: e.g., the ICD-10 for unspecified abdominal pain is R10.9, which was converted to “unspecified abdominal pain”. For this step, hierarchical ICD code mapping was implemented using the “regexp_replace” SQL function, by combining information from the Sub Chapter, Major and Short Description levels, as previously described [13]. These levels match the diagnosis category, name and description respectively, obtained from the most updated (2020) ICD Data R package (http://cran.nexr.com/web/packages/icd/icd.pdf) [13].

In clinical settings, a PI patient might be assigned multiple ICD codes corresponding to general or more specific characterization of PI. To avoid biasing causal modeling, all other ICD codes that were relevant to immunodeficiency were removed as data leaks (Supplementary Table 2) [13].

### Converting ICD codes to phenotypes

We used the PheWAS Phecode v.1.2 system to translate features into clinically meaningful phenotypes (disease categories), prior to BN modeling [21]. One or more ICD codes were mapped into a distinct phenotype across each patient, based on the PheWAS Phecode v.1.2. To perform this, we employed the “regexp_replace” SQL function, combining data from multiple description levels (i.e., the Short and Long Description, Major and Sub Chapter levels), as previously described [13]. This mapping was based on the updated ICD Data R package (http://cran.nexr.com/web/packages/icd/icd.pdf) [13].

### Pre-processing

Following data preparation, the number of clinical history ICD codes identified in Cohorts 1-4 were: 2,188; 2,154; 3,522; and 10,445 ICD codes, respectively. After ICD to phenotype conversions, Cohorts 1-4 involved: 1,590; 1,551; 4,595 and 39,823 phenotypes, respectively. Across all cohorts, BN modeling was performed on phenotype data.

To remove sparse, redundant data and to improve computational efficiency, we performed dimensionality reduction. First, we removed sparse phenotypes that had <5% prevalence in the CID/CVID cases within each cohort. This led to 565, 562, 397 and 331 phenotypes in Cohorts 1-4, respectively. Subsequently, we performed Pearson’s X^2^ analysis to evaluate collinearity between phenotypes within each cohort. Given that all phenotypes were binary and had a hierarchical structure (from general to specific phenotypes), there were many highly colinear phenotype pairs. Based on expert advice from 3 clinical immunologists (co-authors RT, JR and VHT), we only allowed one phenotype from each pair demonstrating a Pearson’s X^2^ statistic P-value < 10^-20^, 10^-20^, 10^-84^ and 10^-84^ in Cohorts 1-4, respectively. This led to 241, 245, 212 and 122 phenotypes in Cohorts 1-4, respectively. Due to the varying selection criteria across cohorts and the large sample size in Cohorts 3 and 4, which increased statistical power, P-value thresholds were chosen to ensure at least 20 phenotypes were included in the DAG across all cohorts [22].

### Causal discovery

Causal discovery aims to recover causal relationships among the variables. Causal networks (CNs), a foundational ML approach rooted in BNs, offer a mathematically rigorous, semantically sound and interpretable representation of cause-effect relationships through probabilistic graphical models that represent variables as nodes and associations as arcs in a DAG [17].

The processes of learning the DAG and the parameters of BNs [23, 24], performing inference and model validation [25], as well as generating hypotheses [26] and guiding the design of experiments (with BNs) [27], are well-studied topics. BNs are generative models: as such, we can use them as a working model of reality and explore the phenomena we are studying through inference, reducing the need for experimental data collection. Furthermore, BNs can easily incorporate information available from the literature and domain experts [28].

To construct his causal reasoning framework, Judea Pearl endowed probabilistic interpretations of BN models with additional causal meaning [16]. Under additional assumptions such as the lack of unobserved (latent) confounders, he showed that we can attribute causal meaning to the BN arcs. Modern literature focuses on how to learn them from observational data [29, 30], from a combination of observational and interventional data [19], and hierarchical data such as that arising from multi-center clinical trials [31]. Further work on BNs has been focused to identify when they can be uniquely identifiable [32], to deal with missing data [33, 34] and to detect possible sources of confounding [35].

Formally, BNs are defined as a set of variables *X*_1_, …, *X*_*N*_ that are associated with the nodes of a DAG *G*. Each arc *X*_*i*_ → *X*_*j*_ indicates that *X*_*i*_ and *X*_*j*_ are linked by a relationship in which *X*_*i*_ is the cause and *X*_*j*_ is the effect. Arcs are assumed not to form cycles in the DAG. Indirect causal effects mediated by other variables are not represented directly as arcs but can be read from the DAG by checking whether *X*_*i*_ and *X*_*j*_ are graphically separated, or if there is an open path that makes it possible to reach *X*_*j*_ from *X*_*i*_.

Each variable has an associated probability distribution. The BN represents the joint probability distributions and provides a clear graphical representation of the relationship among the variables, thus producing an interpretable generative model.

In practice, learning a BN consists of two steps:

1. Learning the structure of the network, i.e., learning which arcs should appear in the DAG to represent the cause-effect relationships between the variables.
2. Learning the parameters of the probability distributions associated with the variables. The BN defines them as the distributions of each variable conditioned on its direct causes, with independent parameters in each distribution.

The first step corresponds to model selection and is the main focus of causal discovery. The second step corresponds to model estimation, a statistical process also integral to causal discovery. Causal discovery and inference were performed using the bnlearn environment (https://www.bnlearn.com/documentation/man/bnlearn-package.html).

### Structure learning

Structure learning involves finding the DAG *G* that is best supported by the data *D*, optimizing for:

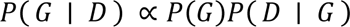

The term *P*(*G*) encodes our prior knowledge on the cause-effect relationships that should appear in the DAG. Further, the likelihood term *P*(*D* ∣ *G*) represents how well the DAG is supported by the data. Together, they are proportional to the posterior probability *P*(*G* ∣ *D*) of the DAG given the data.

Here, we used a score-based approach with tabu search as the causal discovery algorithm and the Bayesian Information Criterion (BIC) to approximate the likelihood of observing the data given the model *P*(*D* ∣ *G*), which was found to provide the best trade-off between speed and structural accuracy [23]. Tabu search is a greedy search algorithm that operates similarly to gradient descent. It chooses to add or remove an arc based on the BIC. BIC is derived as a first-order approximation from *P*(*D* ∣ *G*) and is robust against overfitting.

Furthermore, we employed an ensemble approach by using bootstrapping and model aggregation, to enhance the robustness of our findings by reducing bias and variance across individual DAGs, ultimately identifying the most prevalent variables within the consensus DAGs [36]. We produced 200 bootstrap samples from the data and applied causal discovery to each of them. We then created a “consensus DAG” from the resulting 200 DAGs by selecting those arcs that appeared with a frequency above the data-driven thresholds, as previously detailed [36]. This approach provides us with the inclusion probability of each arc (the frequency with which either *X*_*i*_ → *X*_*j*_ or *X*_*j*_ → *X*_*i*_ appear) and the probability of each causal direction (the frequency of, e.g., *X*_*i*_ → *X*_*j*_ divided by the inclusion probability) for each of the arcs in the consensus BN. These two quantities estimate the posterior probability that *X*_*i*_ and *X*_*j*_ are linked by a cause effect relationship and the possible direction of causality, respectively.

### Parameter Learning

After we have learned the DAG, BNs define the distribution of each variable *X*_*i*_ in the model as *P*(*X*_*i* ∣_ *pa*(*X*_*i*_)), where *pa*(*X*_*i*_) are the direct causes of *X*_*i*_ in the DAG (i.e., all nodes with an arc pointing to *X*_*i*_). As our variables are binary, representing presence or absence of conditions, their distributions are modeled as logistic regressions against their direct causes [16, 17]. The parameters, being regression coefficients, intuitively reflect the odds of causing the associated condition associated with the node [16]. Parameter learning involves estimating these model coefficients, often facilitated by Bayesian inference to incorporate prior knowledge [16, 17].

### Assumptions

Using BNs as CNs requires careful consideration of several essential assumptions. Firstly, inherent to observational studies is the challenge of partial identifiability, where multiple causal models may fit the data equally well, resulting in ambiguity in causal direction [29, 30]. This stems from the inability of observational data alone to differentiate between statistically equivalent models sharing the same dependencies and correlations. Moreover, interpreting arcs as causal effects relies on the assumptions of faithfulness (observed dependencies arise solely from causal structure) and the absence of unobserved confounders [16]. These assumptions, while crucial for valid causal inference, are inherently untestable through statistical methods. Furthermore, the acyclic nature of DAGs precludes representing cyclic relations, which require the construction of dynamic BNs with duplicated nodes across time points, modeled as vector autoregressive series [17, 37, 38]. Lastly, the training data for the BN should be representative, sufficient in quantity (adequate statistical power to identify causal effects), as well as free from sampling bias and systematic missing values which can act as hidden confounders [16, 33]. We observed no missing values in our large-scale diagnosis codes [13].

In our study, to fairly interpret the learned consensus DAGs and evaluate the validity of the aforementioned assumptions, we developed a multi-pronged approach: a) we performed BN model predictions on held-out test data within each cohort and on test data from the other three cohorts (acting as independent datasets); b) we incorporated domain expert knowledge from clinical immunologists to fairly interpret the DAGs across cohorts; c) we performed causal inference by conducting causal interventions on the variables in BN and observing their effects on the odds of being diagnosed with CID/CVID; d) we employed an ensemble approach to enhance the robustness of our findings by reducing bias and variance across individual DAGs, ultimately identifying the most prevalent variables within the consensus DAGs.

### Study-specific assumptions

We set two key study-specific assumptions: 1) that unraveling causal relationships between clinical history phenotypes may improve the identification of CID/CVID (but not the reverse) and 2) that CID/CVID may (commonly) chronologically stem from clinical history phenotypes, given the considerable challenges of underdiagnosis and delayed diagnosis in PI. These assumptions are based on the established association of PI with delayed diagnosis [1, 2, 7–10] and our previous large-scale ML study which demonstrated that clinical history phenotypes consistently preceded the first CID/CVID diagnosis across all four datasets [13]. The latter has also been shown by other computational PI studies [14, 37]. Hence, in our DAG we only allowed the exploration of cause-effect relationships leading from clinical phenotypes towards CID/CVID diagnosis across cohorts (and did not allow the reverse directions). We implement this assumption by prohibiting all the arcs stemming from CID/CVID towards clinical history phenotypes.

### BN model performance

We assessed the predictive performance of our consensus DAGs in two ways:

### Predicting CID/CVID diagnoses within the same population

We used 10-fold cross-validation, training the Bayesian Network (BN) on 9 folds of data and predicting CID/CVID in the held-out fold. This step was repeated across all cohorts.

### Generalizing to different populations

We tested the ability of each consensus DAG to predict CID/CVID in the other three cohorts (using the entire dataset of each cohort). This evaluates the consensus DAG’s ability to generalize to unseen data from distinct populations.

For all evaluations, we perform receiver operating characteristic (ROC) analysis and report the sensitivity, specificity, accuracy, and area under the curve (AUC) as measures of predictive performance.

### Causal inference

We conducted interventional analyses to quantify the impact of each condition (phenotype variable in the DAG) on the odds of receiving a CID and/ or CVID diagnosis (depending on the cohort). We perform an intervention on each phenotype in the consensus DAGs, by removing all incoming arcs and setting its value first to 1 (i.e., a positive diagnosis) and then to 0 (a negative diagnosis). We calculate the odds ratio (OR; presence/ absence of each phenotype) for a positive CID and/CVID diagnosis across phenotypes, to quantify the effect of each condition on the odds of receiving a CID and/ or CVID diagnosis.

By conducting interventions and blocking all incoming causal effects on each phenotype, we can interpret the calculated ORs as cohort-wide causal effects, quantifying how the presence of each phenotype modifies the odds of a CID/CVID diagnosis for each cohort [19].

## Data Availability

The datasets used for this study could not be made publicly available due to a data use commercial agreement between Pfizer and Optum. However, the authors encourage collaborations and would like to declare that the data can be made available to qualified investigators upon request with evidence of institutional review board approval. These data have been previously presented in this publication: https://www.nature.com/articles/s43856-023-00412-8#data-availability. In our current work, we improve the previous methodology for the early identification of these primary immunodeficiencies by developing a novel Causal Bayesian Network methodology.

